# The role of incentives in deciding to receive the available COVID-19 vaccine

**DOI:** 10.1101/2021.08.11.21261829

**Authors:** Liora Shmueli

**Author notes:** **Corresponding author** Liora Shmueli, Ph.D., Department of Management, Bar-Ilan University, Ramat-Gan, 52900, Israel.

## Abstract

**Objective:** To assess the public’s intention to get vaccinated immediately after COVID-19 vaccine became available, and to determine the role of incentives beyond socio-demographic, health-related and behavioral factors, in predicting this intention.

**Methods:** An online survey was conducted among adults in Israel (n=461), immediately after the first COVID-19 vaccine became available (22/12/2020 to 10/1/2021). Two regressions were performed to investigate determinants of intention and sense of urgency to receive the available COVID-19 vaccine.

**Results:** Although many adults intended to receive available COVID-19 vaccine, only 65% intended to immediately receive the vaccine; 16% preferred to wait 3 months and 18% preferred to wait a year. The sense of urgency to get vaccinated differed by age, periphery-level, perceived barriers, cues to action and availability. Monetary rewards or the “green pass” incentives didn’t increase the probability of getting vaccination immediately.

**Conclusions:** Providing data on the role of incentives in increasing the intention to immediately receive the available COVID-19 vaccine is important for health policy makers and healthcare providers. Our findings underscore the importance of COVID-19 vaccine accessibility.

**Practice Implications:** Health policy makers should consider allocating funds for making the vaccine accessible and encourage methods of persuasion, instead of investing funds in monetary incentives.

## 1. Background

On December 31, 2020, the World Health Organization (WHO) issued an Emergency Use Listing (EUL) for the first COVID-19 vaccine, Pfizer-BioNTech, followed by authorizations for other vaccines developed by Moderna, AstraZeneca/Oxford and Jansen. Soon after, several mass vaccination campaigns were initiated around the world [1,2]. Despite their availability, the success of COVID-19 vaccines greatly depends on the proportion of the population that intends to be vaccinated. Specifically, a fraction of the population is not expected to get vaccinated due to the phenomenon known as vaccine hesitancy [3], concerns about the safety of the vaccine or regarding the speed with which new technologies were used to generate the vaccines, uncertainties about how long immunity would last, and vaccine effectiveness against new variants and preventing transmission [4–7].

Several studies investigated the rate of vaccine acceptance before COVID-19 vaccines became available (dates of survey distribution ranged from February until December, 2020). High response rates were reported in Australia (86%) [8], Ecuador (97%), Malaysia (94%), Indonesia (93%) and China (91%) [6]. A study conducted in Europe, involving participants from Denmark, France, Germany, Italy, Portugal, the Netherlands, and the UK, showed a willingness to vaccinate rate of 74% [9]. Lower rates of vaccine acceptance were found among adults in the United States (69%) [10] and Russia (55%) [6].

Over time, yet still before COVID-19 vaccines became available, several surveys and longitudinal studies demonstrated how intentions of getting a COVID-19 vaccine decrease. According to a survey conducted by Ipsos in partnership with the World Economic Forum, of more than 18,000 adults from 15 countries surveyed, COVID-19 vaccination intent decreased in three months, from 77% in August to 73% in October, 2020 [11]. Specifically, intentions to get vaccinated dropped in 10 of 15 countries, with the biggest decreases seen in China (down by 12 points), Australia (down by 9 points), Spain (down by 8 points), and Brazil (down by 7 points). Three longitudinal studies conducted in the US showed a decline in the intention to receive a COVID-19 vaccine from 74% in April to 54% in December, 2020 [12], [13] [14]. Another longitudinal study showed that from March to August, 2020, the resistance to COVID-19 vaccination had increased by 91% in Ireland and 61% in the UK [15].

Yet, at that point in time when COVID-19 vaccines became available, it was still unclear what proportion of the population would get vaccinated and how quickly they would choose to do so. For example, one study conducted in China showed a sharp decline in the intention to get vaccinated immediately once a vaccine became available, from 58.3% in March to 23.0% in December, 2020 [7].

To better cope with vaccine hesitancy, it is important to identify factors associated with vaccine acceptance. Numerous studies examined socio-demographic and health-related factors, and found that significantly higher proportions of males, older individuals (above 55 years of age [9]), those who had received the seasonal influenza vaccine [6,16–18], white and married individuals, those of higher socio-economic status [16], educated respondents [17,18] and individuals considering themselves at higher risk for COVID-19 [5] exhibited higher intentions to get vaccinated. Several recent studies [17,19–22] also added behavioral factors based on the Health Belief Model (HBM), and found that higher levels of perceived benefits from the COVID-19 vaccine, perceived severity of COVID-19 infection, cues to action and trust in the healthcare system or vaccine manufacturers were positively correlated with vaccine acceptance, whereas perceived access, barriers and harm were negatively correlated.

To actively promote voluntary uptake of COVID-19 vaccines, especially among individuals who do not intend to get vaccinated immediately or those who do not intend to get vaccinated at all, several incentive-based strategies were proposed. Some of the proposed strategies included monetary rewards. For example, in the USA, John Delaney and Robert Litan suggested paying people $1000-1500 for vaccination [23,24]. Another strategy suggested by several governments, including those of Chile, Germany, Italy, the UK, and the USA, was the use of “immunity passports” and “vaccination certificates” [25]. Accordingly, the Israeli Ministry of Health developed an incentives model termed the “green pass”. The “green pass” is an entry permit to facilities and social events, such as hotels, restaurants and concerts, for those who recovered from COVID-19 and for fully vaccinated individuals [24,26]. Nevertheless, previous studies did not investigate the role of incentives, beyond socio-demographic, health-related and behavioral factors, in individuals’ willingness to get vaccinated against COVID-19.

As such, the goal of this paper is two-fold, namely to assess the public’s intention to get vaccinated immediately once COVID-19 vaccine became available, and to determine the role of incentives, beyond socio-demographic, health-related and behavioral factors, in predicting this intention.

## 2. Methods

### Study participants and survey design

A cross-sectional national anonymous web-based survey was conducted using an electronic questionnaire, distributed via online social platforms (Google, Facebook and WhatsApp) among the general Israeli adult population (i.e., 18 years old or older). The survey was conducted between December 22, 2020 and January 10, 2021, immediately after the first vaccine became available and mass vaccination campaigns against COVID-19 were initiated in Israel. The questionnaire is partly based on a previous questionnaire distributed to the general public in June, 2020, which was pilot-tested by a panel of experts in the field, including a statistician, a behavioral psychologist and an epidemiologist [17].

### Questionnaire

The questionnaire consisted of the following sections: (1) Socio-demographic predictor variables; (2) health-related predictor variables; (3) HBM predictor variables and (4) incentives for immediate COVID-19 vaccination. Overall, the questionnaire consisted of 36 questions.

### Variables and measurements

The first dependent variable was intention to receive the COVID-19 vaccine, originally measured as a one-item question on a 1-6 scale (1 - not appropriate at all; 6 - very appropriate). This variable was transformed to a binary variable (1 - intend to get vaccinated; 0 - do not intend to get vaccinated) so as to simplify the analyses, allowing us to compare individuals who intend to get vaccinated with those who do not. The second dependent variable was the sense of urgency to receive the COVID-19 vaccine, measured by 3 categories, namely, get vaccinated immediately, get vaccinated within 3 months, and get vaccinated within 1 year.

Independent variables were grouped into four blocks:

1. Socio-demographic predictor variables included: (1) Age group; (2) gender; (3) education level; (4) personal status (in partnership or not; living with or without children); (5) socio-economic level, based on the Israeli Central Bureau of Statistics scale; and (6) periphery level, defined by residential area. The age variable was transformed from numeric to categorical (18-39; 40-59, 60+) in order to address differences between specific age groups.
2. Health-related predictor variables included: (1) Perceived health status; (2) suffering from chronic disease (one or more of the following: Heart disease; vascular disease and/or stroke; diabetes mellitus; hypertension; chronic lung disease, including asthma or immune suppression); (3) smoking; (4) being over-weight; (5) past episodes of COVID-19; (6) past episodes of influenza; and (7) having received influenza vaccine
3. HBM predictor variables included: (1) Perceived susceptibility (included two items); (2) perceived severity (included two items); (3) perceived benefits (included two items); (4) perceived barriers (included one item); (5) cues to action (included five items); and (6) health motivation (included two items). Items in the HBM were measured on a 1-6 scale (1-not appropriate at all; 6 - very appropriate). Negative items were reverse-scored. Scores for each item were averaged to obtain each of the HBM-independent categories. The Cronbach α internal reliability method revealed the internal consistency of the HBM to be Cronbach α=0.796
4. Incentive-related predictor variables included: (1) Availability (“If the vaccine is accessible and available”); (2) monetary reward; (3) “green pass” (“If I receive a “green pass” that will allow various reliefs (entry to places of entertainment etc.)”; and (4) monetary penalty (“If the government cuts my social security benefits or imposes another fine if I do not get vaccinated”).

### Statistical analyses

Data from the electronic questionnaires were imported into SPSS 26 software and identified by code alone. Data processing and analysis was done using SPSS 26 software. To test the reliability of HBM measures, Cronbach’s α test was used. To describe characteristics of the study population, the following methods of descriptive statistics were used: Frequencies, percentages, averages and standard deviations. Relationships between dependent and independent variables were examined by univariate analysis, using either t-tests on independent samples or Chi-squared tests, depending on the characteristics of the variable examined.

Two regressions were performed. First, to investigate determinants of intention to receive COVID-19 vaccine, a four-step hierarchical binary logistic regression was performed. The intention to receive COVID-19 vaccine served as the dependent variable. In the initial step, only socio-demographic variables that were found to have a significant effect (p < .05) on the intention to get vaccinated against COVID-19 were inserted into the regression model as predictors. In the second step, health-related variables that were significant in the univariate analysis were entered as predictors. In the third step, all HBM variables were entered into the model as predictors. In the fourth and final step, four incentive-related variables were entered as predictors (availability, monetary rewards and penalties and “green pass”).

Second, to estimate the sense of urgency to receive COVID-19 vaccine, a multinomial logistic regression was estimated. Specifically, I was interested in predicting what would increase the probability of preferring to get vaccinated within 3 months or within a year from the moment the vaccine became available, rather than getting vaccinated immediately. Only socio-demographic and health-related variables that deemed to have a significant effect (p < .05) in the univariate analysis on the urgency to receive the COVID-19 vaccine were inserted into the regression model as predictors. All HBM and incentive-related variables were entered into the model as predictors.

## 3. Results

### Participant characteristics

Descriptive characteristics of the respondents are provided in Table S1. Overall, 461 respondents completed the survey, 56% of whom were female (n=257). Almost half of the participants were aged 18-39 years. The majority of those included in the study hold an academic degree (n=341), most live with a partner (73%) or with a child (63%). Almost half of the participants were assigned to a middle socio-economic level or live in a locality in the center of the country. Fourteen percent of respondents (n=60) stated that they suffer from at least one chronic disease. Although only 48% had received flu vaccine in the current year (n=221), 42% (n= 195) stated they did not plan to get vaccinated this year.

### Willingness to receive COVID-19 vaccine by the sense of urgency

Overall, 65% of participants were willing to receive the COVID-19 vaccine immediately (n=279) in the week that the vaccine becomes available in Israel, 17% were willing to get vaccinated within 3 months (n=73) and 18% within a year (n=75) from the moment vaccines arrive. Thirty-four participants stated their intention to never get vaccinated, and as such were not included in the sense of urgency to get vaccinated analysis.

### Univariate analyses

#### Intention to receive the COVID-19 vaccine

Results of univariate analyses between socio-demographic and health-related variables and willingness to get vaccinated against COVID-19 are reported in Table S1. Predictor variables that were found to have a statistically significant effect (p<0.05) on the intention to receive COVID-19 vaccine were gender, socio-economic level, suffering from a chronic disease and having received influenza vaccine. Predictor variables that were not found to be statistically significant included age, educational level, personal status, periphery level, being over-weight, smoking, past episodes of COVID-19, past episodes of influenza and perceived health status.

Results of univariate analyses between HBM and incentive-related variables and the intention to get vaccinated against COVID-19 are reported in Table S2. The results in Table S2 indicate that according to HBM, perceived susceptibility, perceived benefits and cues to action were found to have a statistically significant effect (p<0.05) on the intention to receive COVID-19 vaccine. There were no significant differences between the groups in terms of perceived severity, perceived barriers or health motivation. The results in Tables S2 also indicate that according to the incentive-related variables, all four incentives (availability, monetary rewards and penalties and “green pass”) were found to have statistically significant effects (p<0.05) on the intention to receive COVID-19 vaccine.

#### Sense of urgency to receive the COVID-19 vaccine

Results of univariate analyses between socio-demographic and health-related variables and the sense of urgency to receive the COVID-19 vaccine are reported in Table S3. Predictor variables determined to have a statistically significant effect (p<0.05) on the sense of urgency to receive COVID-19 vaccine were age, gender, socio-economic level, periphery level and having received influenza vaccine. Predictor variables not found to be statistically significant included educational level, personal status, suffering from chronic disease, being over-weight, smoking, past episodes of COVID-19, past episodes of influenza and perceived health status.

Results of univariate analyses between HBM variables, incentive-related variables and sense of urgency to receive the COVID-19 vaccine are reported in Table S4. The results in this case are completely consistent with those reported in Table S2 for the case of the intention to receive the COVID-19 vaccine.

### Multivariate analyses

#### Predictors of the intention to receive the available COVID-19 vaccine

The first regression analysis, which included incentive-related beyond socio-demographic, health-related and behavioral factors, explained 76% of the variance in the intention to receive the available COVID-19 vaccine (adjusted R^2^ = 0.76). All model steps were significant. The most important components of the hierarchical regression were the HBM dimensions, which added 48% to the explained variance, on top of the 21% explained by socio-demographic and health-related characteristics. The four incentives added 7% beyond those offered by the HBM.

More specifically, according to the final model, among socio-demographic variables, only gender was associated with the intention to get the available COVID-19 vaccine. Men were more likely to receive the available COVID-19 vaccine than were woman (OR=3.09, 95% CI 1.12–8.53). Only one health-related variable, i.e., having received influenza vaccine, was a significant predictor. Respondents who had not received, and do not plan to receive the seasonal influenza vaccine this winter, are less likely to get vaccinated for COVID-19, as compared with those who received the seasonal influenza vaccine (OR=0.33, 95% CI 0.12–0.91). According to the HBM, perceived benefits (OR=2.50, 95% CI 1.50–4.21) and perceived severity (OR=1.61, 95% CI 1.04–2.49) remained positive significant predictors of vaccine acceptance. Among the incentive-related variables, only perceived availability of the vaccine (OR=2.69, 95% CI 1.80–4.03) was associated with intention to get vaccinated against COVID-19.

A complete description of all model steps, goodness of fit indices and regression coefficients are provided in Table 1.

**Table 1.**
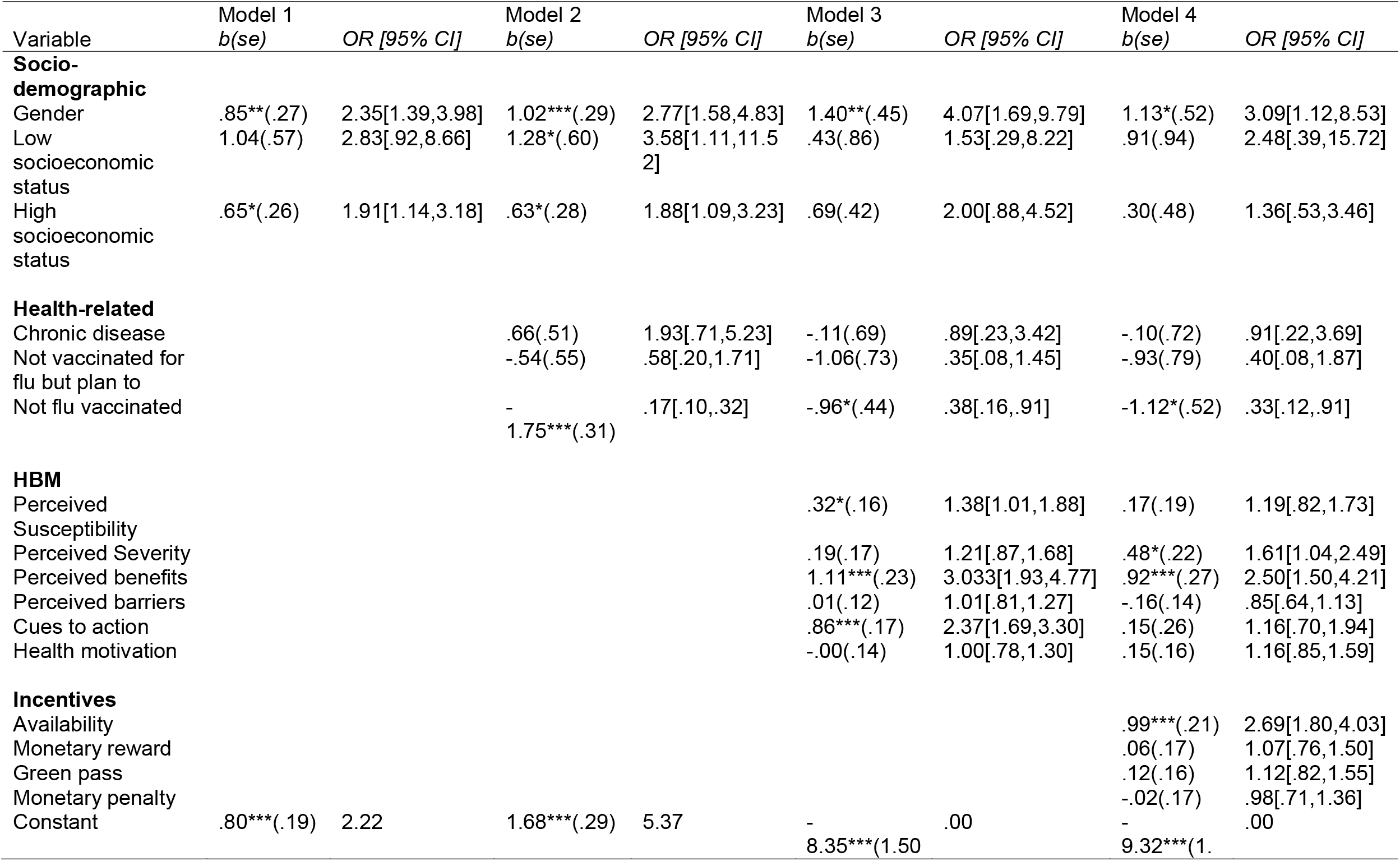
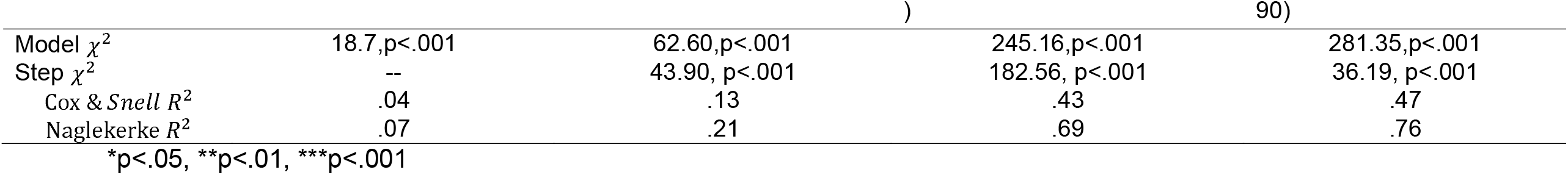
Hierarchical logistic regression analysis - predictors of intention to receive the available COVID-19 vaccine

#### Predictors of sense of urgency to receive the available COVID-19 vaccine

The second regression analyzed the sense of urgency to receive the available COVID-19 vaccine. I was specifically interested in predicting what would increase the intention to get vaccinated immediately, rather than within 3 months or within a year from the time the vaccine became available.

The estimated model was found to be significant. Specifically, according to the model, among socio-demographic variables, age and periphery level were found to be significantly associated with the sense of urgency to receive the current COVID-19 vaccine (p<0.05). Those between the ages of 18-39 years and 40-59 years (OR=4.98, 95% CI 1.23-20.11 and OR=4.18, 95% CI 1.07-16.39, respectively) are over 4 times more likely to get vaccinated in 3 months than to get vaccinated immediately, as compared with people over 60 years of age.

Regarding periphery level, people living in intermediate regions of the country (i.e., between the central urban region and the periphery) are 0.28 times less likely to get vaccinated within a year (OR=0.28, 95% CI 0.09-0.91) than to get vaccinated immediately, as compared to those living in the center of the country.

Among HBM variables, perceived barriers and cues to action were associated with the sense of urgency to receive the available COVID-19 vaccine. For each unit increase in perceived barriers, the likelihood of getting vaccinated in 3 months rather than immediately increased 1.25-fold (OR=1.25, 95% CI 1.03-1.51), while the likelihood of getting vaccinated within a year rather than immediately increased by 1.62-fold (R=1.62, 95% CI 1.20-2.19). It was also found that for each unit increase in perceived cues to action, there was a 40% reduction in the odds of being vaccinated within a year, rather than immediately (OR=0.60, 95% CI 0.37-0.96).

Among the incentive-related variables, only vaccine availability was associated with the sense of urgency to receive the available COVID-19 vaccine. Specifically, for each unit increase in perceived availability, the likelihood of getting vaccinated in 3 months decreased 0.5-fold, while the likelihood of getting vaccinated within a year decreased 0.19-fold, as compared with those who intended to get vaccinated immediately (OR=.50, 95% CI 0.36-0.68 and OR=1.62, 95% CI 0.12-0.31, respectively).

Variables that were not were not found to be significantly associated with the sense of urgency of receiving the available COVID-19 vaccine were: gender, socio-economic level, having received flu vaccine, perceived susceptibility, perceived severity, perceived benefits, and health motivation. Among the incentive-related variables, monetary rewards, monetary penalties and “green pass” were not found to be significant as well.

A complete description of the model, goodness of fit indices and regression coefficients are presented in Table 2.

**Table 2.**
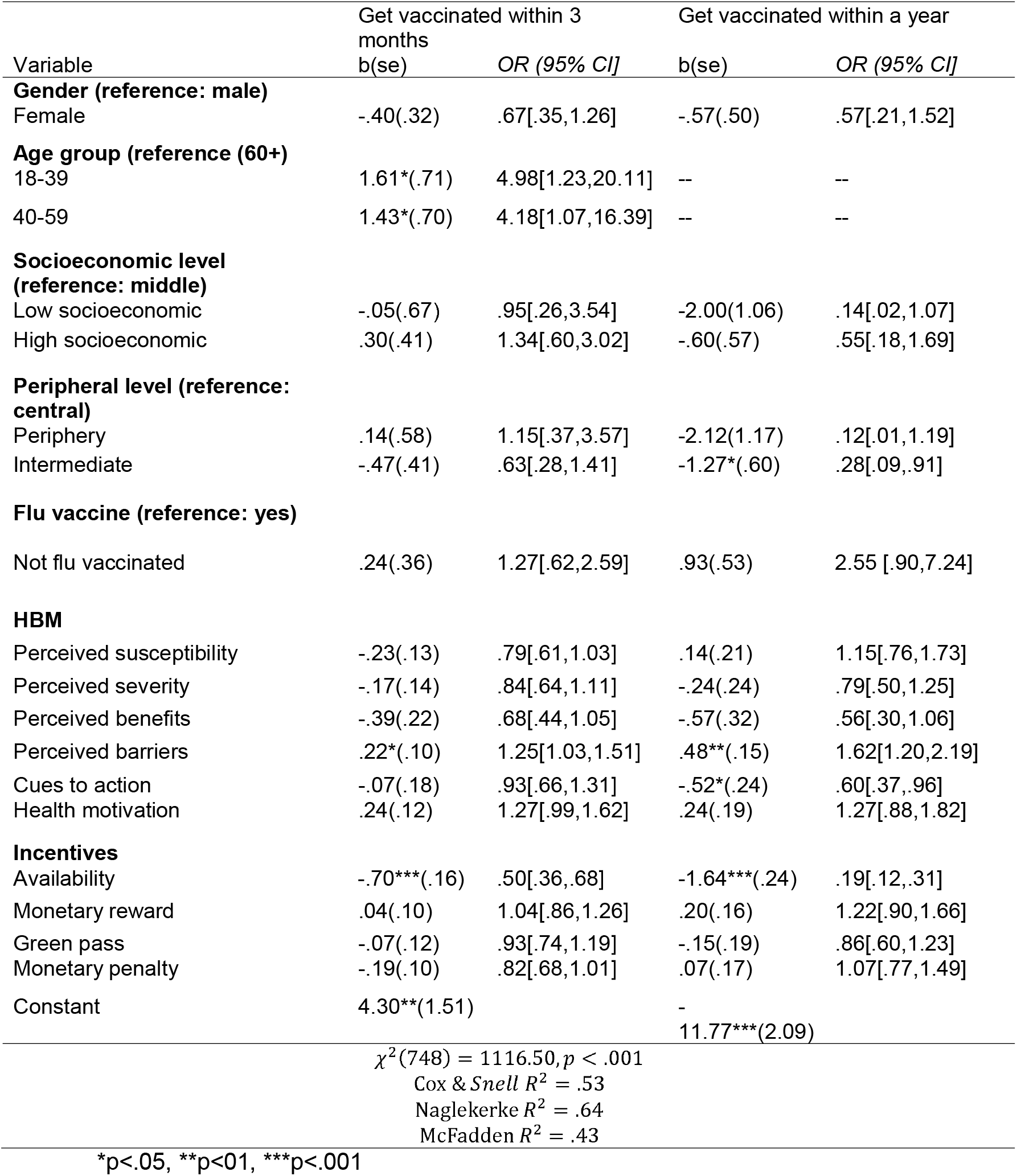
multinomial logistic regression- predictors of sense of urgency to receive the available COVID-19 vaccine

## 4. Discussion and Conclusion

### Discussion

This is apparently the first study to have investigated the role of incentive-related and not only socio-demographic, health-related and behavioral factors in predicting the intention of the general public to immediately receive the available COVID-19 vaccine.

The present study reported the high intention of 81% of those questioned to get vaccinated with the available COVID-19 vaccine during the extensive vaccination campaign held in Israel in late December, 2020, which in the initial phase was characterized by rapid and effective rollout [4]. This finding is consistent with the overall intention to receive the COVID-19 vaccine (80%) found in a previous survey conducted in May, 2020 before the vaccine was available [17]. In both studies, similar socio-demographic, health-related and behavioral predictors of willingness to get vaccinated were found, and it was shown that men and those who reported having been vaccinated against influenza were more likely to receive the COVID-19 vaccine. Moreover, participants were more likely to be willing to get vaccinated if they reported higher levels of perceived benefits and of perceived severity of COVID-19 infection.

However, the present study found that the intention of getting vaccinated immediately with the available COVID-19 vaccine was lower, with only 65% of participants being willing to immediately receive the vaccine. Of the few studies that examined the intention to vaccinate immediately when the vaccine becomes available, some reported even lower rates. Specifically, the study conducted by Mahmud et al. in January, 2021 in Bangladesh showed that only 35.14% were willing to vaccinate immediately [27], while Wang et al. found that in China, the intention of immediate vaccination declined substantially from 58.3% In March, 2020 to 23.0% in November– December, 2020, a bit before the vaccine became available [7].

A large proportion of those who were willing to get vaccinated preferred to wait a period of time, specifically 16% were willing to get vaccinated within 3 months and 18% within a year. A multinomial regression revealed that people between the ages of 18-59 years preferred to get vaccinated within 3 months, as compared with those over 60 years of age, who intended to get vaccinated immediately. This is reasonable as individuals aged 60 years and above are included in the high-risk group for COVID-19. Also, people living in intermediate regions according to a periphery index criterion are less likely to get vaccinated within a year than they are to get vaccinated immediately, as compared with people living the center of the country. One explanation for this may be related to financial considerations, as some of those living far from the center of the country suffered economically during the periods of lockdown.

In this study, several incentives proposed by health policy makers (i.e. monetary rewards, “green pass”, etc.) were added to the predictive model. Among these incentives, only perceived availability of the vaccine was associated with the intention to receive such a vaccine. Neither financial incentives, such as monetary rewards or monetary penalties, nor non-financial incentives, such as the “green pass” were found to be significant predictors of the intention to get vaccinated. Previous studies noted several problems with the use of financial incentives for encouraging COVID-19 vaccination. First, paying people to get vaccinated offends the moral sense of individuals and the community [23], as it is accepted that people have the duty to promote their own health and that the community has a duty to promote their health and social benefits. Second, previous studies demonstrated that monetary incentives do not increase the willingness to get vaccinated against COVID-19, as monetary payment for vaccination is likely to be small and is unlikely to compensate for the risk (perceived or real) of vaccination but only for the inconvenience. Hence, a small payment is unlikely to overcome this view. Nevertheless, larger payments may provide both economic and ethical justifications [28,29]. Third, arguments against financial incentives are related to trust in the pharmaceutical companies producing COVID-19 vaccine and in governmental policymakers [30,31], as offering payment can damage trust in the government. According to Pennings et al., other methods of persuasion, rather than incentives, will be the more effective in increasing vaccination rates. For example, the authors suggested allocating funds to allow primary care providers to address public concerns regarding COVID-19 vaccines [29].

### Limitations

It is important to recognize this study’s limitations when interpreting the reported results. One limitation of this study is that a convenience sample of participants was recruited via an online survey. Although the demographic characteristics of study participants were similar to those of the general Israeli population, this limitation should be considered in interpreting the results of the study, as the sample population does not include those minorities who do not have ready access to online surveys, such as the ultra-Orthodox and Arabs. Moreover, the study used self-reporting of willingness behavior regarding the COVID-19 vaccine, which may be biased, unlike monitoring actual vaccination. Additional limitations include the cross-sectional design of the study and lack of available data on non-respondents.

### Conclusions

This study provides up-to-date survey data on the willingness to receive the available COVID-19 vaccine in the general population of Israel, and the role of incentives in agreeing to immediately receive the vaccine, beyond demographic, health-related and behavioral predictors.

The results presented here highlight that although many adults were willing to receive available COVID-19 vaccine, only 65% of the participants were willing to immediately receive the vaccine; 16% preferred to wait 3 months and 18% preferred to wait a year.

The sense of urgency to get vaccinated differed according to a number of socio-demographic, health-related and behavioral characteristics, including age, periphery level, perceived barriers, cues to action and availability. It was also shown that financial incentives did not increase the probability of getting vaccination immediately, beyond demographic, health-related and behavioral predictors.

In summary, the findings of this study underscore the importance of COVID-19 vaccination accessibility. Health policy makers should consider allocating funds to making vaccine accessible in terms of time and place and also encourage methods of persuasion, rather than investing funds in financial incentives.

### Practice implications

Although this study was conducted in Israel, I believe that most of my findings can be generalized to other countries, as Israel is one of the pioneers in quickly implementing vaccination policies for the entire population. For countries in the process of implementing incentives, it is important to take into account that the incentives offered, be they financial incentives, such as monetary rewards or monetary penalties, or non-financial incentives, such as the “green pass”, were not found to be significant predictors of getting vaccinated with the available vaccine. It is, therefore, important to allocate resources to deal with hesitation by reducing risk perceptions, thereby gaining public trust.

## Data Availability

The datasets generated during the current study are not publicly available but are available from the corresponding author on reasonable request.

## List of abbreviations

HBM: Health Belief Model
WHO: World Health Organization
EULs: Emergency Use Listing

## Declarations

### Ethics approval and consent to participate

The study was approved by the Ethics Committee for Non-clinical Studies at Bar-Ilan University in Israel. The ethics form was signed by the committee head on December 20, 2020. Consent for participating in the study was obtained digitally through Google Forms. Specifically, at the beginning of the questionnaire, participants were asked whether they agree to participate in the research and whether they were 18 years old or older so as to be included in the study. Only participants who answered these two questions positively were allowed to continue with the questionnaire. Participants were also informed that their participation was voluntary and that they had the right to leave at any time without providing any explanation. No incentives were provided to study participants.

### Consent for publication

Not applicable

### Authors’ contributions

LS was responsible for study conception and design, data collection and analysis, and writing the manuscript.

## Acknowledgements

Not applicable

## Author information

Liora Shmueli is a lecturer in the Department of Management, Bar-Ilan University.

