## Supplemental Tables 1-4 for "The role of incentives in deciding to receive the available COVID-19 vaccine"

**Table S1**: Characteristics of respondents by intention to get covid-19 vaccine

| **Sociodemographic** | **All subjects**  **(n=461)** | | **DO NOT intend to get vaccinated against COVID-19**  **N=86 (19%)** | | **Intend to get vaccinated against COVID-19**  **N=375 (81%)** | | $\boldsymbol{\chi}\mathbf{2}$ | **p-value** |
| --- | --- | --- | --- | --- | --- | --- | --- | --- |
|  | N | (%) | N | (%) | N | (%) |  |  |
| **Age group**  18 thru 39 | 216 | (46.9%) | 45 | (20.8%) | 171 | (79.2%) | $\chi2(2)=3.48$ | 0.18 |
| 40 thru 59 | 193 | (41.9%) | 36 | (18.7%) | 157 | (81.3%) |  |  |
| 60+ | 52 | (11.3%) | 5 | (9.6%) | 47 | (90.4%) |  |  |
| **Gender**  Male | 204 | (44.3%) | 25 | (12.3%) | 179 | (87.7%) | $\chi2(1)=9.88$ | 0.002* |
| Female | 257 | (55.7%) | 61 | (23.7%) | 196 | (76.3%) |  |  |
| **Education level**  Non-academic | 120 | (26%) | 27 | (22.5%) | 93 | (77.5%) | $\chi2(1)=1.58$ | 0.209 |
| Academic | 341 | (74%) | 59 | (17.3%) | 282 | (82.7%) |  |  |
| **Personal status-in partnership**  Living with a partner | 336 | (72.9%) | 58 | (17.3%) | 278 | (82.7%) | $\chi2(1)=1.59$ | 0.208 |
| Not living with a partner | 125 | (27.1%) | 28 | (22.4%) | 97 | (77.6%) |  |  |
| **Personal status-with kids**  Living with a child | 290 | (62.9%) | 47 | (16.2%) | 243 | (83.8%) | $\chi2(1)=3.09$ | 0.079 |
| Not living with a child | 171 | (37.1%) | 39 | (22.8%) | 132 | (77.2%) |  |  |
| **Socio-economic level**  Low | 39 | (8.5%) | 4 | (10.3%) | 35 | (89.7%) | $\chi2(2)=7.56$ | 0.023* |
| Middle | 213 | (46.4%) | 51 | (23.9%) | 162 | (76.1%) |  |  |
| High | 207 | (45.1%) | 31 | (15%) | 176 | (85%) |  |  |
| **Peripheral level**  Periphery | 53 | (11.5%) | 6 | (11.3%) | 47 | (88.7%) |  | 0.237 |
| Intermediate | 167 | (36.4%) | 29 | (17.4%) | 138 | (82.6%) | $\chi2\left( 2 \right)=2.88$ |  |
| Central | 239 | (52.1%) | 50 | (20.9%) | 189 | (79.1%) |  |  |

| **Health related variables** | | **All subjects**  **(n=461)** | | | **DO NOT intend to get vaccinated against COVID-19**  **N=86 (19%)** | | | **Intend to get vaccinated against COVID-19**  **N=375 (81%)** | | $\boldsymbol{\chi}\mathbf{2}$ | **p-value** |
| --- | --- | --- | --- | --- | --- | --- | --- | --- | --- | --- | --- |
|  | N | | | (%) | N | (%) | N | | (%) |  |  |
| **Chronic Disease**  No chronic disease | | | 382 | (86.4%) | 78 | (20.4%) | 304 | | (79.6%) | $\chi2\left( 1 \right)=4.97$ | .026* |
| Chronic disease | | | 60 | (13.6%) | 5 | (8.3%) | 55 | | (91.7%) |  |  |
| **Smoking**  Yes | | | 70 | (15.6%) | 12 | (17.1%) | 58 | | (82.9%) | $\chi2\left( 1 \right)=0.07$ | .799 |
| No‎/quitted | | | 380 | (84.4%) | 70 | (18.4%) | 310 | | (81.6%) |  |  |
| **Over weight**  Yes | | | 122 | (27.7%) | 18 | (14.8%) | 104 | | (85.2%) | $\chi2\left( 1 \right)=1.83$ | .177 |
| No | | | 319 | (72.3%) | 65 | (20.4%) | 254 | | (79.6%) |  |  |
| **Past episodes of COVID-19**  Yes | | | 12 | (2.7%) | 3 | (25%) | 9 | | (75%) | $\chi2(1)=0.38$ | .536 |
| No | | | 433 | (97.3%) | 78 | (18%) | 355 | | (82%) |  |  |
| **Past episodes of influenza**  yes | | | 80 | (17.4%) | 16 | (20%) | 64 | | (80%) | $\chi2\left( 1 \right)=0.15$ | .700 |
| no | | | 380 | (82.6%) | 69 | (18.2%) | 311 | | (81.8%) |  |  |
| **Flu vaccine**  No, I don’t plan to get vaccinated this winter | | | 195 | (42.4%) | 64 | (32.8%) | 131 | | (67.2%) | $\chi2\left( 2 \right)=44.75$ | <.001* |
| No, but I plan to get vaccinated this winter | | | 44 | (9.6%) | 5 | (11.4%) | 39 | | (88.6%) |  |  |
| Yes | | | 221 | (48%) | 17 | (7.7%) | 204 | | (92.3%) |  |  |
| **Perceived health status**  Very good | | | 358 | (77.5%) | 67 | (18.7%) | 291 | | (81.3%) | $\chi2(2)=3.58$ | 0.167 |
| Good | | | 90 | (19.5%) | 19 | (21.1%) | 71 | | (78.9%) |  |  |
| Not so good | | | 14 | (3%) | 0 | (0%) | 14 | | (100%) |  |  |

Note: Percentages of “Do not Intend to get vaccinated against COVID-19” and “Intend to get vaccinated against COVID-19” are calculated as valid % per each row (i.e., each row sums up to 100%, without missing values).

**p*<0.05

**Table S3:** Characteristics of respondents by sense of urgency to receive the COVID-19 vaccine

| **Sociodemographic** | **All subjects** | | **Immediately** | | **Within 3 months** | | **Within a year** | | $\boldsymbol{\chi}\mathbf{2}$ | **p-value** |
| --- | --- | --- | --- | --- | --- | --- | --- | --- | --- | --- |
|  | N | (%) | N | (%) | N | (%) | N | (%) |  |  |
| **Age group**  18 thru 39 | 198 | (46.4%) | 120 | (60.6%) | 37 | (18.7%) | 41 | (20.7%) | $\chi2\left( 4 \right)=17.28$ | .002* |
| 40 thru 59 | 179 | (41.9%) | 114 | (63.7%) | 31 | (17.3%) | 34 | 19%) |  |  |
| 60+ | 50 | (11.7%) | 45 | (90%) | 5 | )10%) | 0 | (0%) |  |  |
| **Gender**  Male | 191 | (44.8%) | 139 | (72.8%) | 28 | (14.7%) | 24 | (12.6%) | $\chi2(2)=8.83$ | 0.012* |
| Female | 235 | (55.2%) | 140 | (59.6%) | 44 | (18.7%) | 51 | (21.7%) |  |  |
| **Educational level**  Non-academic | 113 | (26.5%) | 75 | (66.4%) | 15 | (13.3%) | 23 | (20.4%) | $\chi2(2)=2.02$ | 0.364 |
| Academic | 314 | (73.5%) | 204 | (65%) | 58 | (20.5%) | 52 | (16.6%) |  |  |
| **Personal status-partnership**  Living with a partner | 316 | (74.2%) | 200 | (63.3%) | 59 | (18.7%) | 57 | (18%) | $\chi2\left( 2 \right)=3.03$ | 0.192 |
| Not living with a partner | 110 | (25.8%) | 79 | (71.8%) | 13 | (11.8%) | 18 | (16.4%) |  |  |
| **Personal status-living with a child**  Living with a child | 272 | (63.7%) | 176 | (64.7%) | 48 | (17.6%) | 48 | (17.6%) | $\chi2(2)=0.18$ | 0.913 |
| Not living with a child | 155 | (36.3%) | 103 | (66.5%) | 25 | (16.1%) | 27 | (17.4%) |  |  |
| **Socioeconomic level**  Low | 37 | (8.7%) | 28 | (75.7%) | 6 | (16.2%) | 3 | (8.1%) | $\chi2(4)=11.50$ | 0.022* |
| Middle | 193 | (45.4%) | 120 | (62.2%) | 27 | (14%) | 46 | (23.8%) |  |  |
| High | 195 | (45.9%) | 129 | (66.2%) | 40 | (20.5%) | 26 | (13.3%) |  |  |
| **Peripheral level**  Periphery | 49 | (11.6%) | 39 | (79.6%) | 7 | (14.3%) | 3 | (6.1%) |  | 0.041* |
| Intermediate | 153 | (36.1%) | 106 | (69.3%) | 22 | (14.4%) | 25 | (16.3%) | $\chi2\left( 4 \right)=9.97$ |  |
| Central | 222 | (52.4%) | 132 | (59.5%) | 44 | (19.8%) | 46 | (20.7%) |  |  |

| **Health related variables** | **All subjects** | | | **Immediately** | | **Within 3 months** | | **Within a year** | | $\boldsymbol{\chi}\mathbf{2}$ | **p-value** |
| --- | --- | --- | --- | --- | --- | --- | --- | --- | --- | --- | --- |
|  | | N | (%) | N | (%) | N | (%) | N | (%) |  |  |
| **Chronic Disease**  No chronic disease | | 349 | (85.1%) | 219 | (62.8%) | 63 | (18.1%) | 67 | (19.2%) | $\chi2\left( 2 \right)=4.17$ | .13 |
| Chronic disease | | 61 | (14.9%) | 46 | (75.4%) | 9 | (14.8%) | 6 | (9.8%) |  |  |
| **Smoking**  Yes | | 62 | (14.9%) | 47 | (75.8%) | 9 | (14.5%) | 6 | (9.7%) | $\chi2\left( 2 \right)=4.32$ | .115 |
| No‎/quitted | | 354 | (85.1%) | 224 | (63.3%) | 61 | (17.2%) | 69 | (19.5%) |  |  |
| **Over weight**  Yes | | 115 | (28.1%) | 79 | (68.7%) | 20 | (17.4%) | 16 | (13.9%) | $\chi2\left( 2 \right)=1.77$ | .414 |
| No | | 294 | (71.9%) | 186 | (63.3%) | 51 | (17.3%) | 57 | (19.4%) |  |  |
| **Past episodes of COVID-19**  Yes | | 11 | (2.7%) | 6 | (54.5%) | 2 | (18.2%) | 3 | (27.3%) | $\chi2(2)=1.09$ | .579 |
| No | | 403 | (97.3%) | 268 | (66.5%) | 71 | (17.6%) | 64 | (15.9%) |  |  |
| **Past episodes of influenza**  yes | | 74 | (17.4%) | 51 | (68.9%) | 13 | (17.6%) | 10 | (13.5%) | $\chi2\left( 2 \right)=1.05$ | .593 |
| no | | 352 | (82.6%) | 227 | (64.5%) | 60 | (17%) | 65 | (18.5%) |  |  |
| **Flu vaccine**  No, I don’t plan to get vaccinated this winter | | 166 | (39.1%) | 85 | (51.2%) | 33 | (19.9%) | 48 | (28.9% | $\chi2\left( 4 \right)=31.49$ | <.001* |
| No, but I plan to get vaccinated this winter | | 44 | (10.4%) | 28 | (63.6%) | 9 | (20.5%) | 7 | (15.9%) |  |  |
| Yes | | 215 | (50.6%) | 164 | (76.3%) | 31 | (14.4%) | 20 | (9.3%) |  |  |
| **Perceived health status**  Very good | | 327 | (76.6%) | 209 | (63.9%) | 59 | (18%) | 59 | (18%) | $\chi2(4)=2.61$ | 0.626 |
| Good | | 86 | (20.1%) | 60 | (69.8%) | 11 | (12.8%) | 15 | (17.4%) |  |  |
| Not so good | | 14 | (3.3%) | 10 | (71.4%) | 3 | (21.4%) | 1 | (7.1%) |  |  |

Note: Percentages of “Do not Intend to get vaccinated against COVID-19” and “Intend to get vaccinated against COVID-19” are calculated as valid % per each row (i.e., each row sums up to 100%, without missing values).

**p*<0.05

**Table S2**: Univariate analyses between HBM, incentives variables and willingness to get vaccinated against COVID-19

|  | DO not-intend to get vaccinated  (n= 86) | | | Intend to get vaccinated  (n= 375) | | | | | t-test | | P value (two-tail) |
| --- | --- | --- | --- | --- | --- | --- | --- | --- | --- | --- | --- |
| **Variables** | Mean (SD) | | | Mean (SD) | | | | | |  |  |
| **HBM variables** |  | | | |  |  | |  | |  |  |
| Perceived Susceptibility | 2.84 | (1.39) | 5.04 | | | | (1.21) | | -14.67 | | <.001 |
| Perceived Severity | 2.68 | (1.42) | 2.98 | | | | (1.24) | | -1.96 | | .051 |
| Perceived Benefits | 3.40 | (1.32) | 5.38 | | | | (.75) | | -13.39 | | <.001 |
| Perceived Barriers | 3.22 | (1.76) | 3.04 | | | | (1.67) | | .86 | | .39 |
| Cues to action | 2.10 | (1.12) | 4.21 | | | | (1.15) | | -15.41 | | <.001 |
| Health motivation | 4.21 | (1.45) | 4.00 | | | | (1.39) | | 1.23 | | .220 |
| **Incentives variables** |  |  |  | | | |  | |  | |  |
| Availability | 2.09 | (1.14) | 5.31 | | | | (1.21) | | -22.50 | | <.001 |
| Monetary reward | 1.69 | (1.18) | 3.03 | | | | (2.05) | | -5.84 | | <.001 |
| Green pass | 2.88 | (1.88) | 4.91 | | | | (1.59) | | -9.30 | | <.001 |
| Monetary penalty | 2.81 | (1.90) | 3.95 | | | | (1.94) | | -4.93 | | <.001 |

Note: COVID-19 vaccination intention measured by the item: “I want to get vaccinated against the COVID-19 virus now that a vaccine is available”, on a 1-6 agreement scale

HBM and incentives Items Response scale: 1-6 agreement

**Table S4:** Univariate analyses between HBM, incentives variables and the sense of urgency to receive COVID-19 vaccine

|  | Immediately  (n= 279) | | Within 3 months (n= 73) | | Within a year  (n= 75) | | F-test | P value |
| --- | --- | --- | --- | --- | --- | --- | --- | --- |
| **Variables** | Mean | (SD) | Mean | (SD) | Mean | (SD) |  |  |
| **HBM variables** |  |  |  |  |  |  |  |  |
| Perceived Susceptibility | 5.23 | (1.11) | 4.27 | (1.35) | 3.86 | (1.46) | 46.19 | <.001 |
| Perceived Severity | 3.06 | (1.28) | 2.80 | (1.15) | 2.80 | (1.28) | 2.08 | .127 |
| Perceived Benefits | 5.49 | (.72) | 4.97 | (.76) | 4.32 | (1.18) | 63.58 | <.001 |
| Perceived Barriers | 2.96 | (1.68) | 3.32 | (1.65) | 3.30 | (1.70) | 2.13 | .12 |
| Cues to action | 4.41 | (.99) | 3.72 | (1.18) | 2.80 | (1.32) | 67.20 | <.001 |
| Health motivation | 3.90 | (1.41) | 4.23 | (1.39) | 4.11 | (1.31) | 1.84 | .161 |
| **Incentives variables** |  |  |  |  |  |  |  |  |
| Availability | 5.64 | (.88) | 4.61 | (1.28) | 2.84 | (1.42) | 209.45 | <.001 |
| Monetary reward | 3.07 | (2.11) | 2.76 | (1.86) | 2.47 | (1.69) | 2.92 | (.06) |
| Green pass | 5.11 | (1.52) | 4.48 | (1.62) | 3.58 | (1.80) | 28.39 | <.001 |
| Monetary penalty | 4.13 | (1.97) | 3.35 | (1.78) | 3.44 | (1.78) | 7.15 | .001 |

Note: the sense of urgency to receive COVID-19 vaccine measured by the item: "now as the vaccine is available, how soon will you get vaccinated? Immediately, within 3 months or within a year?".

HBM and incentives Items Response scale: 1-6 agreement
